# Direct Oral Anticoagulants Affect Activated Clotting Time During and Bleeding Events After Percutaneous Coronary Intervention

**DOI:** 10.1101/2023.02.28.23286600

**Authors:** Eiji Shibahashi, Takuro Abe, Kazuho Kamishima, Suguru Ebihara, Tetsu Moriyama, Kensuke Shimazaki, Katsumi Saito, Yasuko Uchigata, Kentaro Jujo

## Abstract

**Background:** Inappropriately high activated clotting time (ACT) during percutaneous coronary intervention (PCI) is associated with an increased risk of bleeding events. However, whether the prescription of direct oral anticoagulants (DOAC) affects ACT kinetics during heparin use and adverse clinical events in patients undergoing PCI remains unclear. To evaluate the ACT changes during and adverse clinical events after PCI in patients who were prescribed DOAC.

**Methods:** This observational study included 246 patients undergoing PCI at the two cardiovascular centers who were not receiving warfarin and whose ACT was recorded immediately before and 30 min after injection of unfractionated heparin (UFH). Patients were divided into two groups according to DOAC prescription at the time of the index PCI: DOAC users (n=31) and non-users (n=215). Any bleeding and systemic thromboembolic events were investigated until 30 days after PCI.

**Results:** The average age of this population was 70.5 years, and 66.3% were male. Average ACT was significantly higher in DOAC users than non-users both before and 30 min after UFH induction (157.2 ± 30.1 vs. 131.8 ± 25.1 sec, p<0.001; 371.1 ± 122.2 vs. 308.3 ± 82.2 sec, p<0.001; respectively). The incidence of post-PCI systemic thromboembolism was low and comparable between the two groups (0% vs. 3.7%, p=0.60). However, the rate of any bleeding event was significantly higher in DOAC users than non-users (16.1% vs. 4.7%, p=0.028).

**Conclusion:** Patients receiving DOAC have higher ACTs during PCI and higher incidence of bleeding events than those not receiving DOAC.

**CONDENSED ABSTRACT:** Changes in activated clotting time (ACT) and incidence of systemic thromboembolic and bleeding events in patients undergoing percutaneous coronary intervention (PCI) using conventional heparin were compared between those receiving direct oral anticoagulants (DOAC) and those who were not. ACT both before and 30 min after initial heparin injection was higher in patients who received DOAC than in those who did not. DOAC prescription did not affect the incidence of systemic thromboembolic events. Conversely, patients receiving DOAC more frequently experienced post-PCI bleeding events than those not receiving DOAC.

## INTRODUCTION

Unfractionated heparin (UFH) is a standard intravenous anticoagulant used during percutaneous coronary intervention (PCI), and has been used as a comparator in prior trials of novel anticoagulants.^1-3^ Intravenous UFH may help prevent clot formation on the surfaces of guiding catheters, guidewires, balloons, and metal stents.^4,5^ Previous studies have reported a relationship between activated clotting time (ACT) and thromboembolic complications during PCI.^6-8^ However, a recent meta-analysis revealed inconsistent probabilities of major adverse cardiovascular and bleeding events with various peak ACT values from multiple randomized controlled trials.^9^ The recommended initial dose of UFH is 70-100 U/kg intravenous bolus injection, with additional doses if necessary to achieve the appropriate ACT values.^10^

With the progressively aging society, PCI for patients with atrial fibrillation is being increasingly performed, and combination therapy of anticoagulants and antiplatelet agents has become the standard regimen for these patients.^11^ For anticoagulants, unless contraindicated, direct oral anticoagulants (DOAC) are generally preferred over warfarin, because DOAC have comparable efficacy in preventing thromboembolic events and cause fewer bleeding events.^12-15^ However, the appropriate initial UFH dose during PCI for patients receiving DOAC remains unclear. The guideline-recommended initial UFH dose for patients not receiving DOAC may be excessive for those receiving them, leading to a possible increase in bleeding events. Therefore, we aimed to evaluate the effect of DOAC on the kinetics of ACT during PCI and periprocedural thrombotic and bleeding events with the usual UFH dosage, comparing patients receiving DOAC and those who were not.

## METHODS

### Study population, procedural steps, and endpoints

This observational cohort study initially included 654 consecutive patients who underwent PCI at two cardiovascular centers in Japan between February 2017 and April 2018. However, patients who received warfarin or for whom ACT was not measured during PCI were excluded. This study was registered in the University Hospital Medical Information Network Clinical Trials Registry (UMIN-CTR No. UMIN000047650). ACT kinetics during and bleeding events after PCI were compared between DOAC users and non-users. In both facilities, to ensure efficacy and safety, 100 U/kg of UFH was routinely administered as the initial dose, and ACT was measured immediately before and 30 min after UFH administration using a Hemochron device (Soma Tech. Inc., Bloomfield, CT, USA). If ACT was < 250 sec at 30 min after initial UFH, an additional dose was intravenously administered to achieve an ACT > 250-300 sec, as per guidelines.^10^ An additional UFH dose of 40 U/kg is stipulated by the guidelines; however, the target ACT and actual dose of additional UFH were left to the discretion of the operators. To investigate the relationship between ACT values during PCI and bleeding events, bleeding events were recorded for up to 1 month after discharge. Bleeding within 72 h after PCI or before discharge was evaluated and defined as a periprocedural bleeding event.^16^ Bleeding events were classified into major and minor bleeding events according to the International Society on Thrombosis and Haemostasis criteria.^17^ Major bleeding was defined as bleeding in which at least one of the following was observed: (i) fatal bleeding; (ii) symptomatic bleeding in a critical organ or space, such as intracranial, intraspinal, intraocular, retroperitoneal, intra-articular, pericardial, or intramuscular with compartment syndrome; and (iii) bleeding causing ≥ 2.0 g/dL (1.24 mmol/L) decrease in the hemoglobin level, or leading to transfusion of two or more units of whole blood or red blood cells. To evaluate thromboembolic events, occurrence of ischemic stroke, myocardial infarction, acute stent thrombosis, or other thromboses was investigated for up to 1 month after discharge. This study was conducted in accordance with the principles of the Declaration of Helsinki. The ethics committee of our institution approved the study protocol (Kosei Hospital: March 7, 2022, Nishiarai Heart Center Hospital: January 14, 2022) and waived the requirement for written informed consent because of the retrospective study design.

### Data collection and follow-up

We evaluated the baseline characteristics of the patients, including age, sex, body mass index, history of smoking and revascularization therapy, and other comorbidities; blood count (platelets and hemoglobin level); blood chemistry test (serum albumin, serum creatinine, estimated glomerular filtration rate (eGFR), lipid profiles, including low- and high-density lipoprotein cholesterols and triglyceride, and glycated hemoglobin); blood coagulation tests (prothrombin time-international normalized ratio [PT-INR] and activated partial thromboplastin time); clinical presentations of coronary artery disease (ST- and non-ST-segment elevation myocardial infarctions, unstable angina pectoris, and chronic coronary syndrome [CCS]); and procedural parameters for PCI (approach site, sheath size). For the radial approach, a 5-7 Fr sheath dedicated to the radial artery was used (Radifocus Introducer II ®, Terumo, Tokyo, Japan) and for the femoral approach, a 5-8 Fr sheath dedicated to the femoral artery (Radifocus Introducer II ®, Terumo, Tokyo, Japan). Oral medications, including aspirin, thienopyridines, ticagrelor, cilostazol, DOAC, beta blockers, angiotensin-converting enzyme inhibitors or angiotensin II receptor blockers, calcium channel blockers, diuretics, nitrates, proton pump inhibitors, and statins, being taken at the time of PCI were recorded.

### Statistical analysis

Data are expressed as means and standard deviations and numbers and percentages. For continuous variables, the independent Student’s t-test and non-parametric equivalent Mann– Whitney U test were used to compare the two groups. Fisher’s exact test was used to evaluate categorical variables. Consecutive samples from DOAC users and non-users were compared using the Holm–Sidak method for multiple t-tests. To evaluate whether inappropriately elevated ACT > 350 sec was associated with bleeding events independent of other known bleeding prognostic factors,^18^ we performed multivariate logistic regression analysis using age (> 75 years), DOAC prescription, severe renal dysfunction (eGFR < 30 mL/min/1.73 m^2^), decreased hemoglobin level (< 11 g/dL), and thrombocytopenia (< 100,000/μL). Statistical significance was set at two-tailed p < 0.05. Statistical analyses were performed using the R software, version 3.3.0 (R Foundation for Statistical Computing, Vienna, Austria).

## RESULTS

### Patient profiles

Of 654 patients, 31 (12.6%) who did and 215 (87.4%) who did not receive DOAC during PCI were analyzed, with the remaining 408 excluded because of a lack of appropriate ACT data or warfarin prescription (**Figure 1**). Baseline patient profiles are listed in **Table 1**. The average age of this population was 70.5 ± 12.6 years, and 66.3% were male. When comparing DOAC users and non-users, atrial fibrillation was significantly more prevalent in DOAC users (87.1% vs. 3.3%, p < 0.001). Although both groups were generally comparable with regard to the prevalence of clinical history, DOAC users showed a significantly higher prevalence of cerebral infarction (25.8% vs. 10.7%, p = 0.04), heart failure (32.3% vs. 15.8%, p = 0.04), and chronic obstructive pulmonary disease (9.7% vs. 0.9%, p = 0.02). In addition, non-DOAC users were more likely to have diabetes, dyslipidemia, and undergo hemodialysis, although the difference was not statistically significant. Of the patients who underwent PCI, 190 (77.2%) were diagnosed with CCS and 179 (72.8%) underwent transradial PCI. In most cases, 6 Fr sheaths were used for vascular access. In the blood tests, PT-INR was higher in DOAC users than in non-users; however, none of the parameters were statistically significant. As for medications, diuretics were more commonly prescribed for DOAC users (54/8% vs. 26.0%, p = 0.003). The prescription rate of prasugrel was significantly higher for DOAC non-users and that of clopidogrel tended to be higher for DOAC users (prasugrel: 54.8% vs. 81.9%, p = 0.002; clopidogrel: 29.0% vs. 14.4%, p = 0.06). However, the prescription rates of dual antiplatelet drugs (DAPT) at the time of PCI did not significantly differ between the two groups.

**Table 1.**
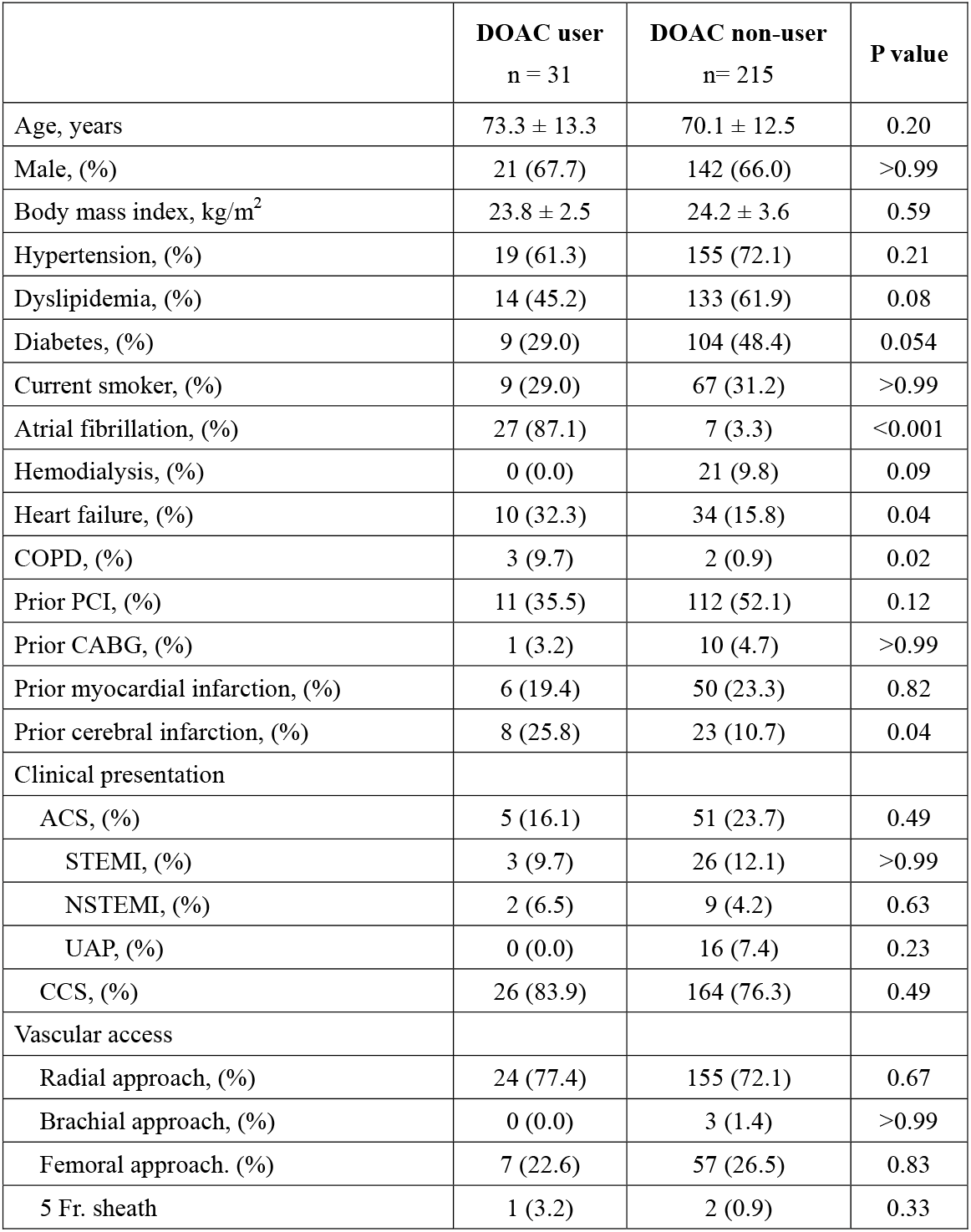
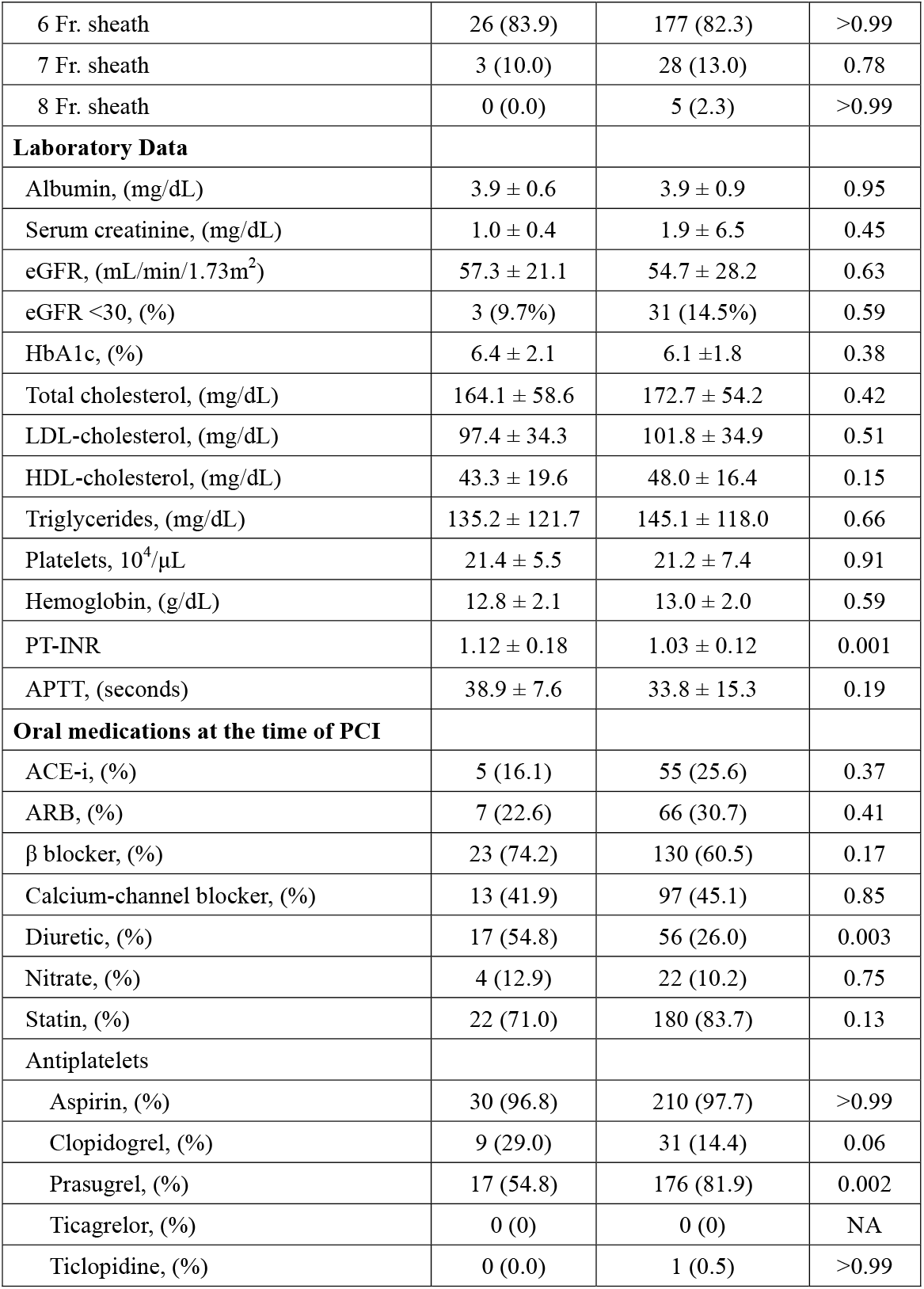
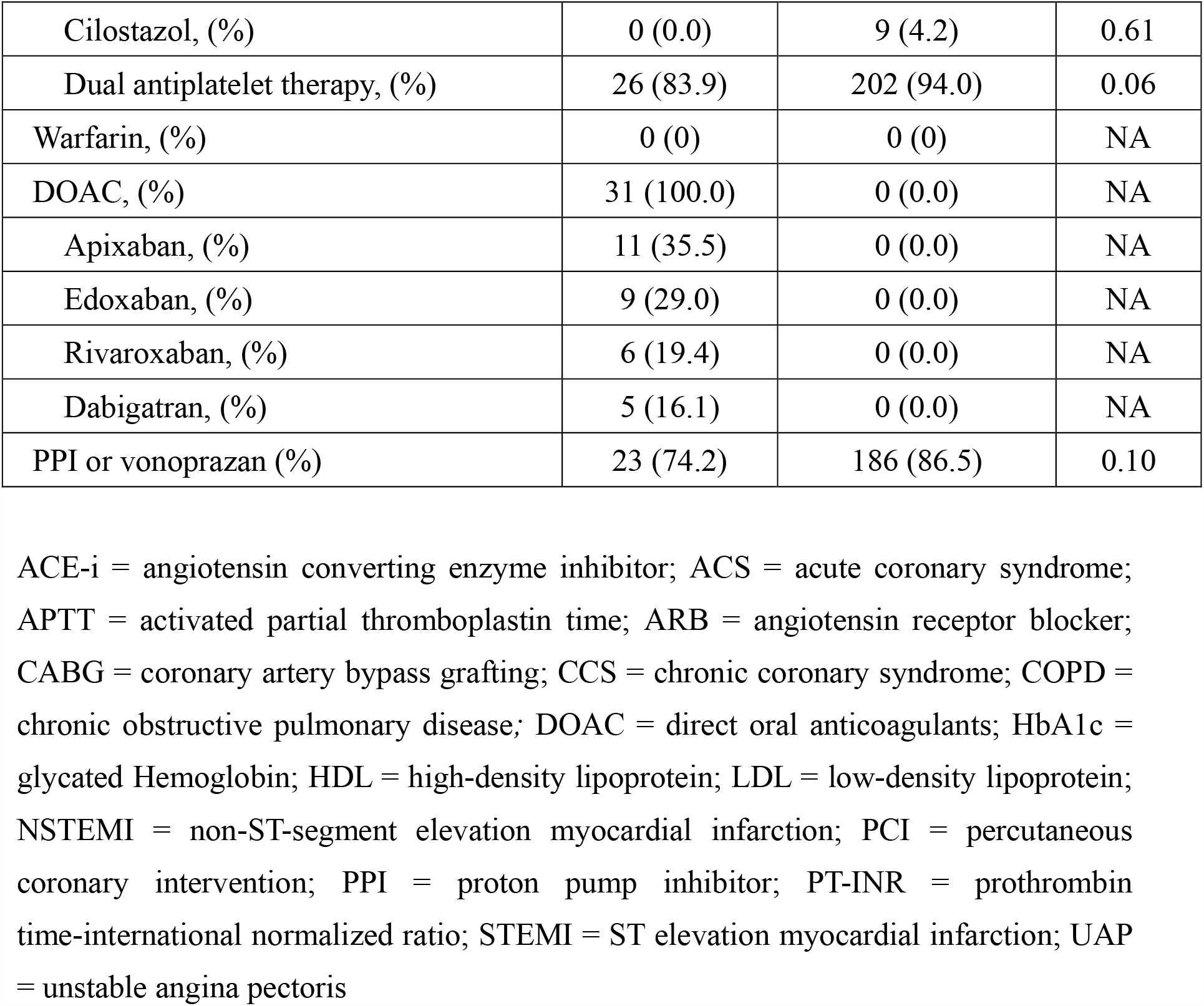
Baseline clinical profiles.

|  | **DOAC user**<br>n = 31 | **DOAC non-user**<br>n= 215 | **P value** |
| --- | --- | --- | --- |
| Age, years | 73.3 ± 13.3 | 70.1 ± 12.5 | 0.20 |
| Male, (%) | 21 (67.7) | 142 (66.0) | >0.99 |
| Body mass index, kg/m<sup>2</sup> | 23.8 ± 2.5 | 24.2 ± 3.6 | 0.59 |
| Hypertension, (%) | 19 (61.3) | 155 (72.1) | 0.21 |
| Dyslipidemia, (%) | 14 (45.2) | 133 (61.9) | 0.08 |
| Diabetes, (%) | 9 (29.0) | 104 (48.4) | 0.054 |
| Current smoker, (%) | 9 (29.0) | 67 (31.2) | >0.99 |
| Atrial fibrillation, (%) | 27 (87.1) | 7 (3.3) | <0.001 |
| Hemodialysis, (%) | 0 (0.0) | 21 (9.8) | 0.09 |
| Heart failure, (%) | 10 (32.3) | 34 (15.8) | 0.04 |
| COPD, (%) | 3 (9.7) | 2 (0.9) | 0.02 |
| Prior PCI, (%) | 11 (35.5) | 112 (52.1) | 0.12 |
| Prior CABG, (%) | 1 (3.2) | 10 (4.7) | >0.99 |
| Prior myocardial infarction, (%) | 6 (19.4) | 50 (23.3) | 0.82 |
| Prior cerebral infarction, (%) | 8 (25.8) | 23 (10.7) | 0.04 |
| Clinical presentation |  |  |  |
| ACS, (%) | 5 (16.1) | 51 (23.7) | 0.49 |
| STEMI, (%) | 3 (9.7) | 26 (12.1) | >0.99 |
| NSTEMI, (%) | 2 (6.5) | 9 (4.2) | 0.63 |
| UAP, (%) | 0 (0.0) | 16 (7.4) | 0.23 |
| CCS, (%) | 26 (83.9) | 164 (76.3) | 0.49 |
| Vascular access |  |  |  |
| Radial approach, (%) | 24 (77.4) | 155 (72.1) | 0.67 |
| Brachial approach, (%) | 0 (0.0) | 3 (1.4) | >0.99 |
| Femoral approach. (%) | 7 (22.6) | 57 (26.5) | 0.83 |
| 5 Fr. sheath | 1 (3.2) | 2 (0.9) | 0.33 |
| 6 Fr. sheath | 26 (83.9) | 177 (82.3) | >0.99 |
| 7 Fr. sheath | 3 (10.0) | 28 (13.0) | 0.78 |
| 8 Fr. sheath | 0 (0.0) | 5 (2.3) | >0.99 |
| **Laboratory Data** |  |  |  |
| Albumin, (mg/dL) | 3.9 ± 0.6 | 3.9 ± 0.9 | 0.95 |
| Serum creatinine, (mg/dL) | 1.0 ± 0.4 | 1.9 ± 6.5 | 0.45 |
| eGFR, (mL/min/1.73m<sup>2</sup>) | 57.3 ± 21.1 | 54.7 ± 28.2 | 0.63 |
| eGFR <30, (%) | 3 (9.7%) | 31 (14.5%) | 0.59 |
| HbA1c, (%) | 6.4 ± 2.1 | 6.1 ± 1.8 | 0.38 |
| Total cholesterol, (mg/dL) | 164.1 ± 58.6 | 172.7 ± 54.2 | 0.42 |
| LDL-cholesterol, (mg/dL) | 97.4 ± 34.3 | 101.8 ± 34.9 | 0.51 |
| HDL-cholesterol, (mg/dL) | 43.3 ± 19.6 | 48.0 ± 16.4 | 0.15 |
| Triglycerides, (mg/dL) | 135.2 ± 121.7 | 145.1 ± 118.0 | 0.66 |
| Platelets, 10<sup>4</sup>/μL | 21.4 ± 5.5 | 21.2 ± 7.4 | 0.91 |
| Hemoglobin, (g/dL) | 12.8 ± 2.1 | 13.0 ± 2.0 | 0.59 |
| PT-INR | 1.12 ± 0.18 | 1.03 ± 0.12 | 0.001 |
| APTT, (seconds) | 38.9 ± 7.6 | 33.8 ± 15.3 | 0.19 |
| **Oral medications at the time of PCI** |  |  |  |
| ACE-i, (%) | 5 (16.1) | 55 (25.6) | 0.37 |
| ARB, (%) | 7 (22.6) | 66 (30.7) | 0.41 |
| β blocker, (%) | 23 (74.2) | 130 (60.5) | 0.17 |
| Calcium-channel blocker, (%) | 13 (41.9) | 97 (45.1) | 0.85 |
| Diuretic, (%) | 17 (54.8) | 56 (26.0) | 0.003 |
| Nitrate, (%) | 4 (12.9) | 22 (10.2) | 0.75 |
| Statin, (%) | 22 (71.0) | 180 (83.7) | 0.13 |
| **Antiplatelets** |  |  |  |
| Aspirin, (%) | 30 (96.8) | 210 (97.7) | >0.99 |
| Clopidogrel, (%) | 9 (29.0) | 31 (14.4) | 0.06 |
| Prasugrel, (%) | 17 (54.8) | 176 (81.9) | 0.002 |
| Ticagrelor, (%) | 0 (0) | 0 (0) | NA |
| Ticlopidine, (%) | 0 (0.0) | 1 (0.5) | >0.99 |
| Cilostazol, (%) | 0 (0.0) | 9 (4.2) | 0.61 |
| Dual antiplatelet therapy, (%) | 26 (83.9) | 202 (94.0) | 0.06 |
| Warfarin, (%) | 0 (0) | 0 (0) | NA |
| DOAC, (%) | 31 (100.0) | 0 (0.0) | NA |
| Apixaban, (%) | 11 (35.5) | 0 (0.0) | NA |
| Edoxaban, (%) | 9 (29.0) | 0 (0.0) | NA |
| Rivaroxaban, (%) | 6 (19.4) | 0 (0.0) | NA |
| Dabigatran, (%) | 5 (16.1) | 0 (0.0) | NA |
| PPI or vonoprazan (%) | 23 (74.2) | 186 (86.5) | 0.10 |

ACE-i = angiotensin converting enzyme inhibitor; ACS = acute coronary syndrome; APTT = activated partial thromboplastin time; ARB = angiotensin receptor blocker; CABG = coronary artery bypass grafting; CCS = chronic coronary syndrome; COPD = chronic obstructive pulmonary disease; DOAC = direct oral anticoagulants; HbA1c = glycated Hemoglobin; HDL = high-density lipoprotein; LDL = low-density lipoprotein; NSTEMI = non-ST-segment elevation myocardial infarction; PCI = percutaneous coronary intervention; PPI = proton pump inhibitor; PT-INR = prothrombin time-international normalized ratio; STEMI = ST elevation myocardial infarction; UAP = unstable angina pectoris

**Figure 1.**
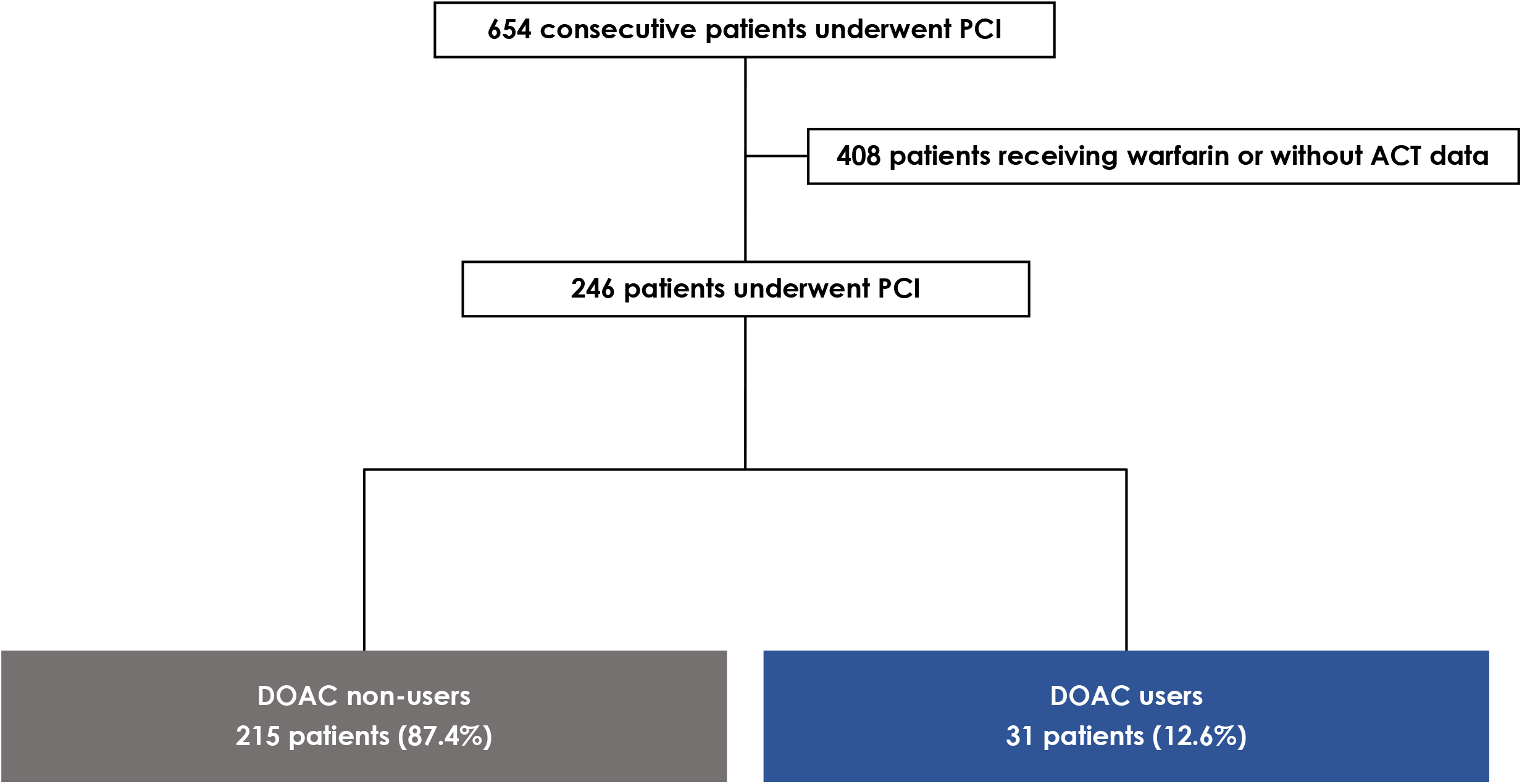
Study population. ACT, activated clotting time; DOAC, direct oral anticoagulants; PCI, percutaneous coronary intervention

### Clinical endpoints

ACT values before and 30 min after UFH administration were significantly higher in DOAC users than in non-users (baseline: 157.2 ± 30.1 vs. 131.8 ± 25.1 sec, p < 0.001; 30 min after UFH: 371.1 ± 122.2 vs. 308.3 ± 82.2 sec, p < 0.001) (**Table 2, Figure 2A**). Additionally, the absolute increase in ACT from baseline to 30 min after UFH was significantly greater in DOAC users than in non-users (ΔACT: 213.9 ± 120.4 vs. 176.5 ± 86.8 sec, p = 0.03) (**Figure 2B**).

**Table 2.** Changes in ACT during PCI.

|  | **DOAC user** | **DOAC non-user** | **P value** |
| --- | --- | --- | --- |
|  | n = 31 | n = 215 |  |
| **Parameters** |  |  |  |
| Baseline ACT, sec | 157.2 ± 30.1 | 131.8 ± 25.1 | <0.001 |
| ACT after 30 minutes of heparin administration, sec | 371.1 ± 122.2 | 308.3 ± 82.2 | <0.001 |
| ΔACT, sec<sup>a</sup> | 213.9 ± 120.4 | 176.5 ± 86.8 | 0.03 |

ACT = activated clotting time; DOAC = direct oral anticoagulant; PCI = percutaneous coronary intervention.
a. ΔACT: (baseline ACT) – (ACT after 30 minutes of heparin administration)

**Figure 2.**
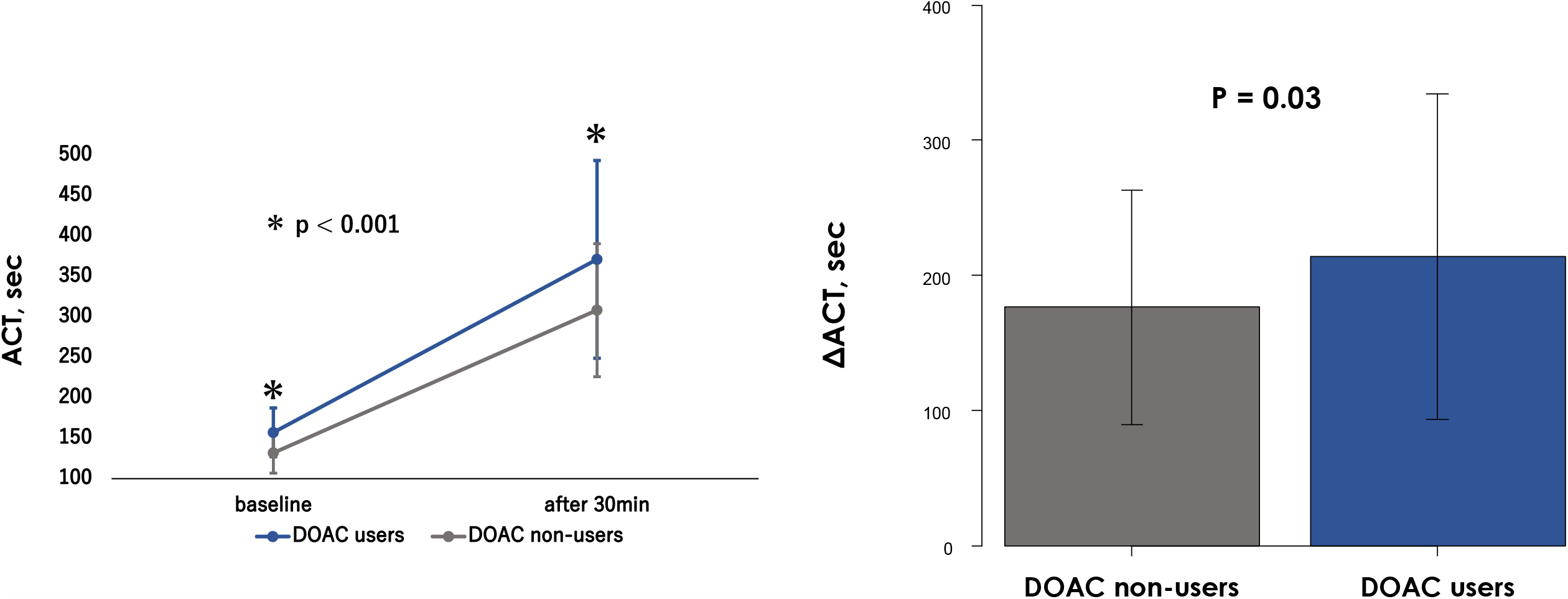
ACT kinetics in DOAC users and non-users. ACT values in DOAC users (blue) and non-users (gray) at baseline and 30 min after heparin administration (A) and ΔACT* (B) are shown. ACT, activated clotting time; DOAC, direct oral anticoagulants * ΔACT: (baseline ACT) – (ACT after 30 min of heparin administration)

Bleeding events were more frequent in DOAC users than in non-users (16.1% vs. 4.7%, p = 0.028) (**Figure 3**). However, none of the bleeding components significantly differed between the two populations (major bleeding: 6.5% vs. 1.4%, p = 0.12; minor bleeding: 9.7% vs. 3.3%, p = 0.12) (**Figure 3**). On the other hand, periprocedural bleeding events in the early postoperative period within 72 h or before discharge were significantly more prevalent in DOAC users than in non-users (12.9% vs. 1.9%, p = 0.01). Conversely, no significant difference was observed between the two groups in terms of thromboembolic events, including ischemic stroke, myocardial infarction, and stent thrombosis (0% vs. 3.7%, p = 0.60) (**Table 3**).

**Table 3.** Clinical outcomes.

| **Outcomes** | **DOAC users** | **DOAC non-user** | **P value** |
| --- | --- | --- | --- |
|  | n = 31 | n = 215 |  |
| Any bleeding, (%) | 5 (16.1) | 10 (4.7) | 0.028 |
| Major bleeding, (%) | 2 (6.5) | 3 (1.4) | 0.12 |
| Minor bleeding, (%) | 3 (9.7) | 7 (3.3) | 0.12 |
| Periprocedural bleeding events*, (%) | 4 (12.9) | 4 (1.9) | 0.010 |
| Bleeding events after 72 hours of PCI | 1 (3.2%) | 6 (2.8%) | >0.99 |
| Ischemic stroke /myocardial infarction/acute stent thrombosis/other thrombosis, (%) | 0 (0) | 8 (3.7%) | 0.60 |
| Ischemic stroke, (%) | 0 (0) | 5 (2.3) | >0.99 |
| Myocardial infarction, (%) | 0 (0) | 1 (0.5) | >0.99 |
| Acute stent thrombosis, (%) | 0 (0) | 1 (0.5) | >0.99 |
| Other thrombosis, (%) | 0 (0) | 1 (0.5) | >0.99 |

DOAC = direct oral anticoagulant; PCI = percutaneous coronary intervention
\* Periprocedural bleeding events; bleeding within 72 hours after PCI or before discharge

**Figure 3.**
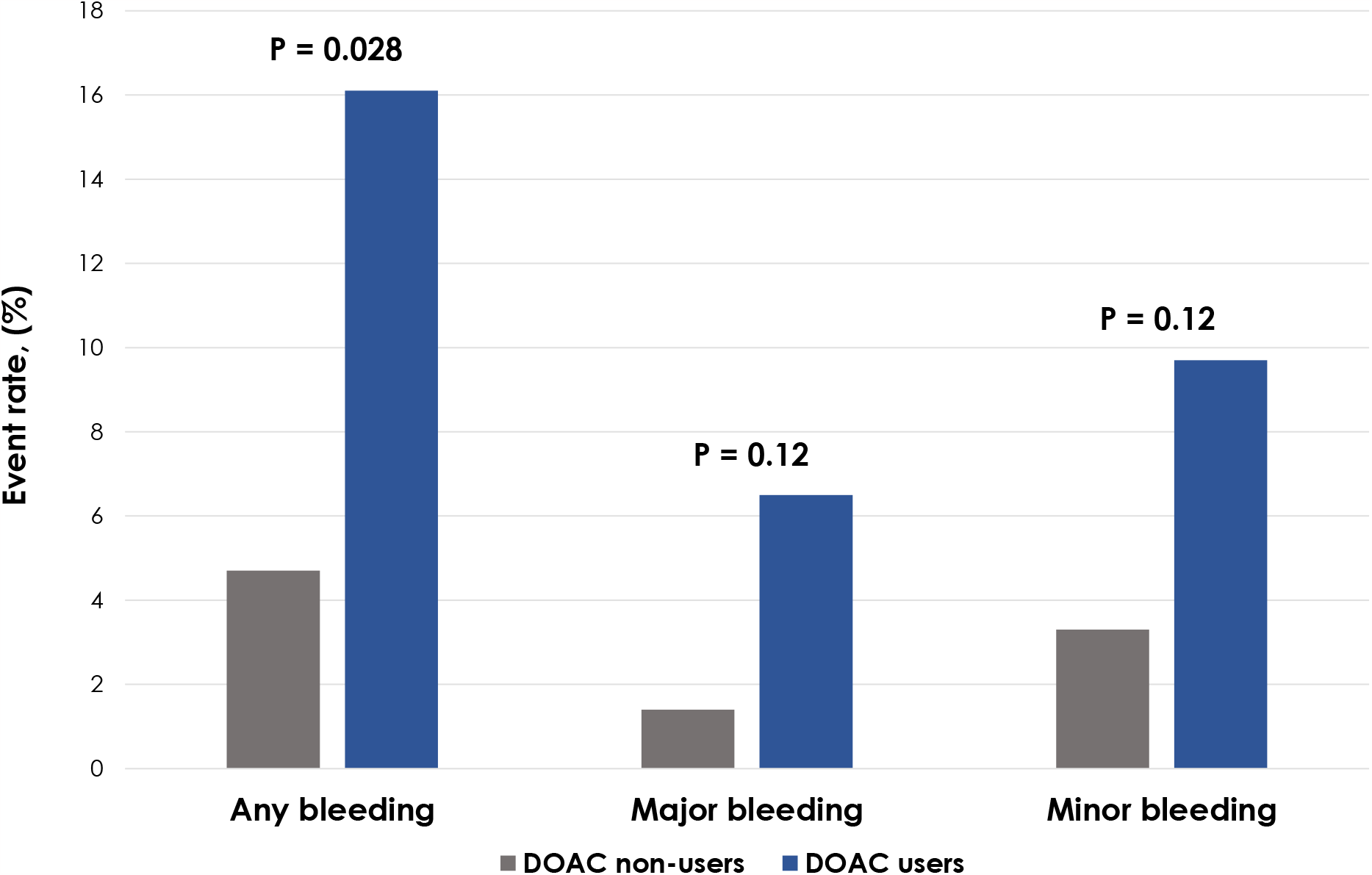
Comparison of any bleeding between DOAC users and non-users. Incidence of any bleeding in DOAC users (blue) and non-users (gray) is shown. DOAC, direct oral anticoagulants

Univariate analysis revealed that excessively high ACT (> 350 sec) was associated with any bleeding event (odds ratio [OR], 4.18; 95% confidence interval [CI], 1.43–12.2; p < 0.01). Multivariate analysis confirmed that elevated ACT, administration of DOAC, and thrombocytopenia were independent predictors of any bleeding (odds ratio [OR]: 3.9, 95% confidence interval [CI]: 1.22–12.5, p = 0.02; OR: 3.75, 95% CI: 1.05–13.4, p = 0.042; OR: 25.1, 95% CI: 2.35–268.0, p < 0.01, respectively) (**Supplemental Table 1**).

## DISCUSSION

To the best of our knowledge, ACT kinetics during and bleeding complications after PCI in patients receiving DOAC have not been investigated. We found that ACTs at baseline and 30 min after guideline-based UFH administration were prolonged during PCI in patients receiving DOAC compared with those who were not. DOAC users had more bleeding events than non-users; however, the occurrence of thromboembolic events did not significantly differ between the two.

### Effect of DOAC on ACT variation

UFH exerts its anticoagulant effect by binding to the enzyme inhibitor antithrombin III, producing a conformational change that increases the antithrombin III-associated deactivation of thrombin, factor Xa, and factor IX.^19^ Therefore, concomitant use of UFH with a DOAC that inhibits thrombin or factor Xa increases, and may even exceed, the intended anticoagulant effect. Moreover, the rapid effect of intravenous anticoagulants with a bolus dose of UFH is unpredictable and influenced by several factors, including age, body weight, sex, blood antithrombin III levels, and platelet count.^20-22^ Owing to these uncertainties, ACT monitoring of the coagulation capacity is necessary. However, the guidelines do not state an appropriate initial UFH dose for patients prescribed DOAC; they receive the same initial bolus dose as those not receiving DOAC.^10^ In the present study, the following two ACT variations were observed in patients receiving DOAC. First, the baseline ACT values before UFH administration were already higher in patients receiving DOAC than in those not receiving them. Therefore, ACT values after UFH administration were > 350 sec in patients receiving DOAC and within the target range of 300-350 sec in those not receiving DOAC. Second, DOAC users had a greater increase in ACT after the first UFH dose than the non-users. These results suggest that the initial UFH dose for DOAC users needs to be reduced from that recommended by current guidelines to achieve the optimal ACT range. Furthermore, the amount of additional UFH should be reconsidered because of the greater ACT increase after UFH administration in patients receiving DOAC.

### Bleeding events

In this study, DOAC users had a higher frequency of bleeding events within 30 days after PCI. Multivariate analysis confirmed that DOAC use was a risk factor for any bleeding at 30 days. In addition, bleeding within 72 h, as a short-term periprocedural complication, was more common in patients receiving DOAC. Periprocedural bleeding also may be related to DOAC use. Severe renal dysfunction and a history of hemodialysis are reportedly associated with increased incidence of bleeding events after PCI.^18^ In this study, the baseline eGFR values and prevalence of severe renal dysfunction did not significantly differ between the two populations. In contrast, DOAC non-users were more likely to undergo regular hemodialysis, although this difference was not significant (hemodialysis: 0% vs. 9.8%, p = 0.09). When adjusted for known risk factors of post-PCI bleeding events, elevated ACT at 30 min after initial bolus administration of UFH was an independent determinant of bleeding events, along with DOAC administration and thrombocytopenia. As for possible preventive interventions, the results of this study suggest that when performing PCI in patients receiving DOAC, a reduced initial bolus dose of UFH may be required to reduce bleeding events. However, this study did not include a protocol with a reduced initial UFH dose for patients receiving DOAC; thus, the safety and efficacy of a potential reduced dose regimen remain unknown. Future trials are required to develop a UFH administration protocol for patients receiving DOAC.

### Thrombotic events

A previous meta-analysis reported that a prolonged ACT of 300-350 sec is associated with fewer thrombotic events in patients not receiving DOAC.^24^ However, the meta-analysis is old, and devices, such as stents and catheters, are outdated. Additionally, recent reports recommending DAPT, including potent P2Y12 inhibitors, have not found a clear relationship between major bleeding, death, or myocardial infarction and ACT.^25^ The optimal ACT value using contemporary devices and DAPT therapy remains uncertain. In this study, although no patient received ticagrelor, they were on a strict DAPT regimen, including P2Y12 inhibitors, and the stent and catheter devices employed were the latest available at the time. Additionally, although the ACT at 30 min after initial UFH administration was prolonged in patients receiving DOAC, there was no significant difference in the occurrence of thrombotic events compared with patients not receiving DOAC. This may be due to the low incidence of thrombotic events in the overall study population. Therefore, we could not confirm whether ACT prolongation was responsible for the reduction in thrombotic events, as reported in recent PCI trials.^26,27^ Furthermore, our study mainly included patients with CCS, and those with acute coronary syndromes (ACS) comprised only a small population (CCS 77.2% vs. ACS 22.8%); catheter and stent thrombosis are common in ACS. Therefore, the efficacy of ACT prolongation in preventing thrombotic events during PCI in patients with ACS receiving DOAC remains controversial.

### Limitations

This was an observational study with a small sample size and limited observation period. Therefore, comparing different pathologies, such as ACS and CCS, and generalizing the results among all patients undergoing PCI are difficult. ACT was measured before and 30 min after UFH administration; however, ACT was not subsequently measured in many patients. Hence, it is unclear how many patients were able to reach the optimal ACT target range by the end of the PCI. In addition, the clinical background associated with bleeding complications was not adjusted, which may have impacted the incidence of bleeding events. In addition, we did not consider the time between DOAC prescription and UFH induction. Furthermore, DAPT duration and implementation were decided by the cardiologist, and ticagrelor was not administered because it was not permitted for patients undergoing PCI in Japan during the study period. Therefore, there was a bias regarding antiplatelet medication, which may have influenced bleeding events. Finally, this study did not include patients requiring the use of anticoagulants other than UFH;^10^ thus, bleeding events due to the use of non-UFH anticoagulants could not be assessed.

## CONCLUSIONS

Conventional UFH administration at the time of PCI results in higher baseline and prolonged ACT values and a greater incidence of post-PCI bleeding events in patients receiving DOAC compared with those who are not. Reducing the initial bolus dose of UFH can be considered to decrease bleeding events when PCI is performed for DOAC users, who are a high-risk population for bleeding.

## Data Availability

All data generated or analyzed during this study are included in this article. Further enquiries can be directed to the corresponding author.

## Acknowledgements

We would like to thank Editage (www.editage.jp) for English language editing.

## ABBREVIATIONS

ACT: activated clotting time
ACS: acute coronary syndrome
CCS: chronic coronary syndrome
DAPT: dual antiplatelet drugs
DOAC: direct oral anticoagulants
PCI: percutaneous coronary intervention
PT-INR: prothrombin time-international normalized ratio
UFH: unfractionated heparin

